# Pathogenicity assessment of a TSC2 c.2966+1G>A splice site variant in vaginal adenocarcinoma

**DOI:** 10.64898/2025.12.31.25343109

**Authors:** Paraic A. Kenny

## Abstract

Clinical sequencing of a vaginal adenocarcinoma specimen yielded a reported TSC2 variant which was classified as splice-modulating loss-of-function, prompting consideration of a clinical trial for tumors with TSC2 loss. Literature review suggested that germline alterations affecting this particular exon are considered non-pathogenic, raising concern about the initial somatic variant interpretation. To resolve this ambiguity, raw data were provided by Tempus and an integrated DNA & RNA analysis allowed detailed characterization of the impact of this alteration on splicing. We confirm that this variant results in the complete exclusion of exon 26 from the transcript, however the exon 25-27 splice results in an in-frame transcript. Analysis of many additional wild-type tumor and normal tissue samples demonstrate that exon 26 is commonly spliced out of TSC2 transcripts, indicating that the transcript containing the exon 25-27 splice is highly unlikely to be pathogenic. This analysis did not support the classification of this variant as pathogenic and suggests that patients with such mutations should not be candidates for therapeutics designed for TSC2 dysfunction.

## INTRODUCTION

TSC2 is a tumor suppressor gene which encodes tuberin. Tuberin interacts with TSC1-encoded hamartin and this complex negatively regulates mTORC1. Heterozygous germline loss of function mutations in either TSC1 or TSC2 cause the autosomal dominant disorder, tuberous sclerosis complex (Henske et al., 2016). As loss of TSC1 or TSC2 function is predicted to elevate mTOR pathway activity, mTOR inhibitors are being evaluated for patient care (e.g. NCT05103358). Accordingly, it is especially important to distinguish passenger from driver variants in these genes for appropriate patient selection.

A specimen from a late 50’s female patient with advanced vaginal adenocarcinoma underwent comprehensive genomic profiling (somatic DNA and RNA analysis, Tempus xT assay) which revealed an alteration in TSC2 (c.2966+1G>A), categorized by Tempus as a “Splice region variant – Loss of Function”. The variant’s high allele frequency (69.4%) was consistent with the possibility of LOH at this locus. This variant was not annotated in either ClinVar or the cBioPortal when this case was initially reviewed in 2022. Currently, this nucleotide variant interpreted discordantly in key data sources for precision oncology. It is rated as a variant of uncertain significance in Clinvar (Accession: VCV001422070.7) and three variants affecting this splice site are rated as pathogenic in cBioportal (Reviewed 12/28/2025).

In considering whether the patient might meet molecular eligibility for a clinical trial targeting TSC2 dysfunction (NCT05103358) we evaluated whether available data were consistent with this mutation being pathogenic. This single base substitution alters the G of the GT splice donor dinucleotide immediately following exon 26 of transcript NM_000548.5. *(Note that this exon is frequently referred to as “Exon 25” in the tuberous sclerosis germline literature in which the non-coding first exon is not counted)*. This exon is known to be spliced out of several transcripts, resulting in an in-frame joining of the immediately upstream and downstream exons, indicating that alterations directly affecting splicing of this exon do not result in a coding sequence frameshift. In considering the likely pathogenicity of any somatic TSC2 alterations, it is important to incorporate the considerable experience of clinical geneticists in evaluating TSC2 variants. Studies of germline alterations affecting this particular exon have concluded that (1) TSC2 transcripts lacking this exon encode TSC2 isoforms that are in-frame and do not deleteriously affect TSC complex function in experimental systems and (2) such variants are not associated with clinically diagnosable tuberous sclerosis (Ekong et al., 2016).

Because this particular variant has not been previously functionally characterized, we retrieved raw data from Tempus to allow an integrated DNA and RNA analysis of TSC2 transcription in this specimen to determine the consequences, if any, of this alteration on the repertoire of transcripts produced by this locus and their possible pathogenicity.

## METHODS

### Tissue specimen and initial clinical NGS analysis

The analyzed tissue was a core needle biopsy from an FDG-avid lung nodule from a patient with recurrent vaginal adenocarcinoma. The specimen was sent to Tempus for comprehensive genomic profiling of both DNA and RNA using the xT assay (Beaubier et al., 2019). For this specimen, tumor content was estimated at 50% and the TSC2 variant was detected at a variant allele frequency of 69.4%.

### NGS data re-analysis

FASTQ and BAM files were provided by Tempus from both the DNA and RNA analyses of the tumor specimen. Analysis was performed by visualization using the Integrated Genomics Viewer (Thorvaldsdottir et al., 2013). The DNA BAM file was analyzed without modification. The RNA BAM file was noted to have been aligned using a non-splice-aware aligner which prevented visualization of splice junctions. To enable splicing analysis we used hisat2 (Kim et al., 2019) was used to generate a new BAM file from the paired FASTQ RNA-Seq reads. The reference genome was hg19.

To directly compare the splicing at the TSC2 locus in this case with other cancer cases sequenced by Tempus on the same platform, we used a convenience sample of 95 cancer cases spanning a range of solid tumor types from the Gundersen Cancer Biobank that had previously been sequenced by Tempus. For this analysis we used the depth tool from samtools (Li et al., 2009) to compute the read depth at the midpoint of each of exons 25, 26 and 27 of Tempus in each sample. All read depths were normalized to the depth at the midpoint of exon 25 for that sample.

### Determining the TSC2 exon 26 splicing pattern in normal tissues

We obtained data from GTEx (GTEx Consortium, 2013) for this purpose, specifically RNA-Seq analysis of 17,382 specimens from 948 individuals, spanning 54 tissue types (dbGAP accession phs000424.v9). Exon-exon junction read counts (for TSC2 exons 25-26, exons 25-27, exons 26-27) were obtained from the following file: GTEx_Analysis_2017-06-05_v8_STARv2.5.3a_junctions.gct. For each specimen, the percentage of total TSC2 reads in which exon 26 was spliced out was calculated.

### Ethical approval

This case is part of the Gundersen Precision Oncology Cohort, a protocol approved by the Institutional Review Board of Gundersen Clinic, Ltd (PI: Kenny; IRB # 2-22-02-003).

## RESULTS

Analysis of RNA-Seq reads spanning splice junctions indicated that exon 26 was completely spliced out in this specimen, as evidenced by the red splicing arcs (Fig 1 A & B, top track) skipping exon 26. In contrast, a representative analysis of a set of five TSC2 wild-type cases showed a mixture of transcripts, some including and some excluding exon 26 (Fig1 A & B, lower tracks).

**Figure 1.**
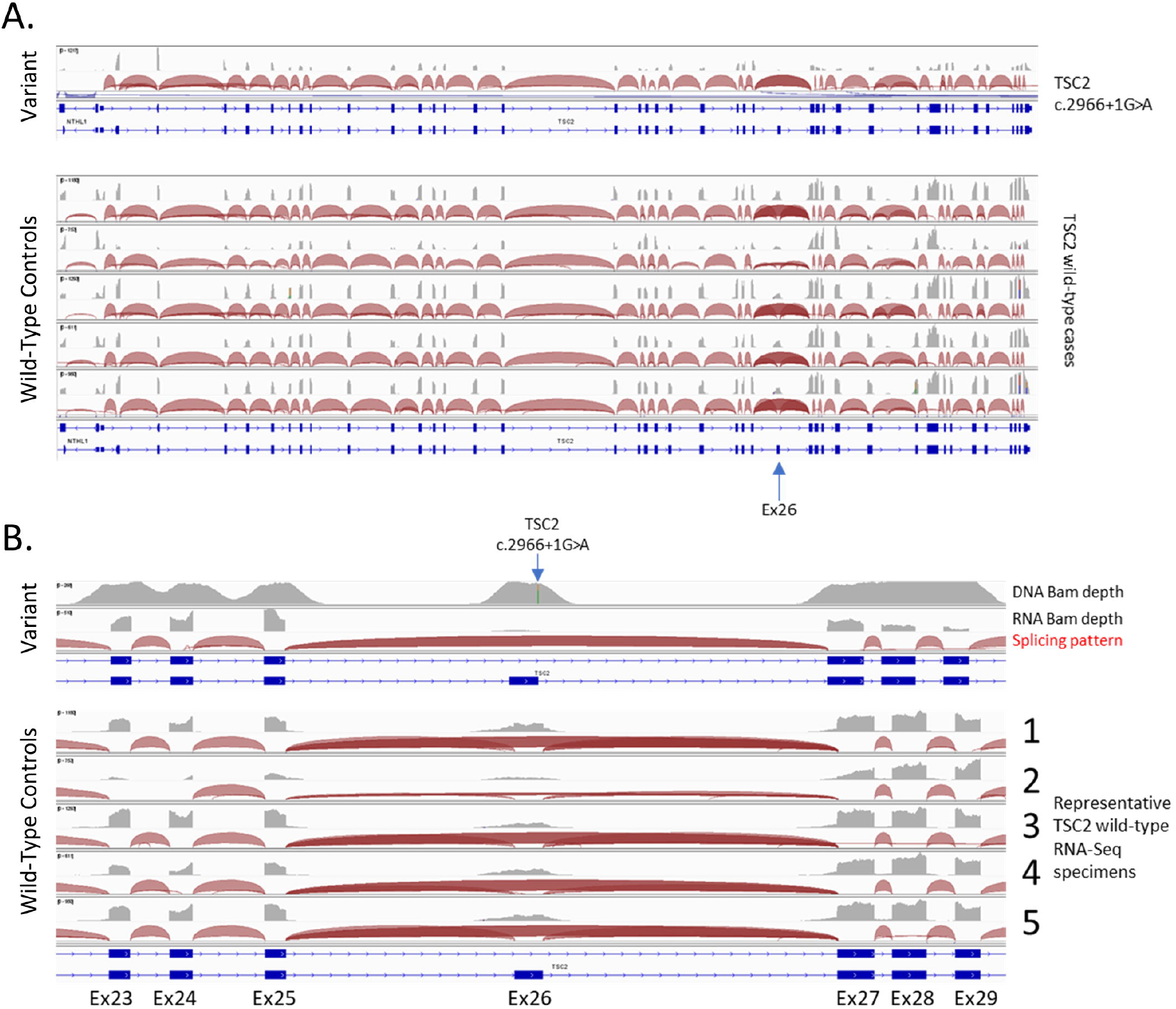
RNA-Sequencing analysis of TSC2 splice patterns in TSC2 c.2966+1G>A case and five TSC2 wild-type controls. (A)Whole-locus overview of TSC2 splicing patterns. (B)TSC2 splicing patterns in the region surrounding exon 26. The mutation in this case can be seen in the DNA BAM histogram at the top of this panel.

In the specimen from the TSC2 splice site mutant sample, the RNA sequencing read depth at Exon 26 was substantially lower than the neighboring exons 25 and 27 and no evidence was detected of any split read involving the junctions of either exons 25-26 or exons 26-27, implying that exon 26 is not included in transcripts from the mutant TSC2 locus.

Several RefSeq transcripts for TSC2 have been reported, which differ by their inclusion of exon 26. To ascertain whether the almost complete absence of exon 26 containing transcripts in this vaginal adenocarcinoma case was unusual, we compared read depth at the mid-points of exons 25, 26, and 27 in this case with a convenience sample of 95 RNA-Seq bam files from cancer cases tested at Tempus (Figure 2). Data shown are normalized to the read depth at the mid-point of exon 25 in each specimen. In most cases the depth at exon 26 is lower than the corresponding depth at exon 25 (implying that exon 26 is spliced out in a proportion of transcripts). This TSC2-mutant case (red) had the lowest relative depth at exon 26 compared to all other specimens. As noted above, the few reads aligning to exon 26 in this specimen lacked any indication of a split mapping to other exons and, in fact, extended into the intronic sequences on either side of the exon suggesting that they are spurious genomic DNA derived reads. Accordingly, it seems that mRNA transcripts in this TSC2 mutant specimen likely have an even lower proportion of exon 26 containing reads than depicted in the Figure.

**Figure 2.**
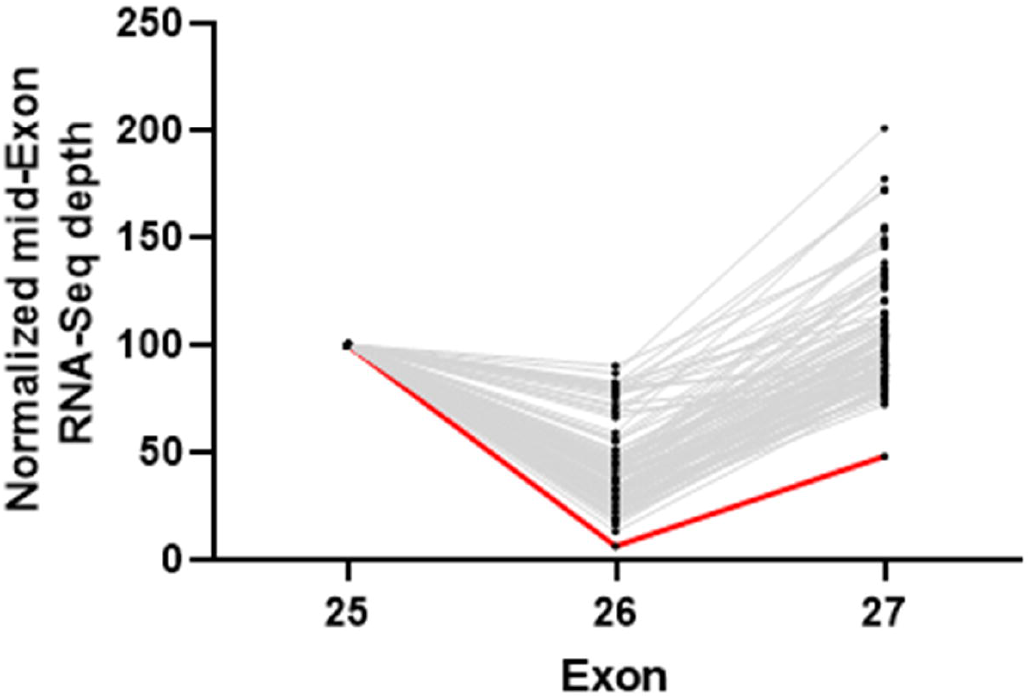
Analysis of relative sequencing depth at TSC2 exons 25, 26 and 27 in the TSC2 c.2966+1G>A case and 95 control cases. All read depths were normalized to the mid-point of exon 25 for each case. The TSC2 c.2966+1G>A case (red) and a control cohort of 95 additional cases sequenced on the Tempus xT platform are shown.

To determine more broadly the frequency at which this exon is skipped in physiologically normal tissues, we evaluated exon junction reads from the GTEx RNA-Seq analysis of 17,382 specimens from 948 individuals, spanning 54 tissue types (dbGAP accession phs000424.v9). After excluding a small number of specimens with fewer than 5 junction spanning reads in this region (n = 193) we found that exon 26 skipping was highly prevalent in all specimens in this dataset. The median proportion of transcripts lacking this exon was 90% (min 17%, LQ 81%, UQ 96% max 100%) suggesting that a mutation that causes this exon to be skipped is highly unlikely to be pathogenic.

## CONCLUSION

The TSC2 c.2966+1G>A splice region variant results in the elimination of transcripts containing exon 26. In all cancer and normal tissue specimens examined, transcripts lacking inclusion of this exon were highly prevalent. Considering also clinical data suggesting that multiple alterations (including frameshifts) affecting this exon are not pathogenic and experimental data suggesting that inclusion of this exon is dispensable for TSC complex function (Ekong et al., 2016), it seems most likely that this somatic mutation (and likely other mutations affecting this exon) are unlikely to be deleterious and that consideration of therapies targeted at TSC2 deficiency is unwarranted for such patients.

## Data Availability

All data produced in the present study are available upon reasonable request to the authors.

## ACKNOWLEDGEMENTS

This study was funded by the Gundersen Medical Foundation. PK holds the Dr. Jon & Betty Kabara Endowed Chair in Precision Oncology.

